# Professionalism and associated factors among nurses working in Hadiya zone public hospitals, Southern Ethiopia. A Facility based cross sectional study (quantitative and qualitative method)

**DOI:** 10.64898/2026.08.21.26360983

**Authors:** Tadele Mengesha Wondafirash, Beriso Furo Wengoro, Ephrem Mamuye Haile, Mitiku Abera Beyene

## Abstract

**Introduction:** Professionalism is a conceptualization of obligations, attributes, interactions, attitudes, and role behaviors required for professionals with individual clients and to the society as a whole. Despite there is a known effect of low professionalism of nurses on health care delivery system, the study determining professionalism and affecting factors in Ethiopia is limited in to a few numbers. Therefore this study was aimed to assess perceived level of professionalism and associated factors among nurses working in Hadiya Zone public hospitals, south Ethiopia.

**Methods:** An institution based concurrent mixed cross sectional study was conducted from May 09 to June 09/2022. A total of **376** nurses were selected by using Computer generated Simple random sampling techniques for quantitative study whereas Purposive sampling technique was used to select qualitative study participants. Epi data version 4.6 was used to enter the data whereas Statistical package for social science version 25 was used for the analysis of quantitative data. Ordinal logistic regression model was used to identify effects of independents on outcome variable.

**Results:** A total of 360 nurses participated in the study with a response rate of 95.7%. The overall low perceived professionalism among nurses working in Hadiya zone found to be 40.3%. Variables such as Educational status (AOR= 0.045 (0.021, 0.096), Sense of accomplishment (AOR= 3.21(1.85, 5.55), and participating in professional associations (AOR= 5.302(2.82, 9.96) were significantly associated with professionalism. Qualitative study finding showed Salary, training, upgrading education; undermining nursing profession, inadequate facilities were the raised influencing factors of professionalism.

**Conclusion:** This study revealed that professionalism among nurses was low and the associated factors were; level of education, taking training, participating in professional organizations, and sense of accomplishment. Ethiopian nursing association and Ministry of Health should focus on developing nurses’ professionalism.

## Introduction

Professionalism is a general concept raised by different communities; which is defined as a conceptualization of obligations, attributes, interactions, attitudes, and role behaviors required for professionals with individual clients and to society as a whole or it is the conduct or qualities that define a profession or a professional person (1, 2). Professionalism in nursing referred to as the integrated belief, ideas and values for nursing and nurses as a profession. Professionalism is more than simply mastering and using technical skills. It is about showing an unwavering commitment to the vocation and the willingness to continuously deliver the highest-quality care to patients (3).

Nurses who value professionalism showed adherence with technical competence and practice standards which is important to have value-based foundations for professionals with ethical commitment for patients’ safety and good outcomes (4). As identified in different studies, a low level of professionalism results the negative outcomes by decreasing productivity, increasing turnover and attrition these can affects delivering qualified care, patient satisfaction, public image toward profession, and health-related indicators (4–6). Criticism in nurses’ skills and abilities also resulted from a lack of professionalism, because it makes nurses to lose confidence to carry out their duties based on scientific evidence, and fail to support the development of the occupation (7).

When nurses exhibit professional behaviors, patients receive better care, team communication improves, there is more accountability among all practitioners, and the clinical atmosphere as a whole is more favorable. As a result, nurses are urged to take part in studies, develop theories, and participate in professional organizations and political activities in order to display professional attitudes (8). But the report of the conducted study revealed that the mean professionalism scores of nurses’ autonomy, membership in professional associations, and written scientific articles are low (5, 9, 10).

The study carried out to determine the professional behavior of nurses in Turkish nurses revealed that, the professionalism levels for nurses were low in the areas of autonomy, publication, and research. Finally, their study concluded that the level of professionalism among studied nurses was low (5).

In Ethiopia, the study that assessed professionalism and its predictors among 332 nurses working in Jimma public hospitals, identified a low level of professionalism among nurses (11). Changes in nursing practice, Lack of autonomy, Lack of leadership skills, working long hours, emotional load and undervalue by society, and Limited opportunities are some major challenges in Nursing Professionalism (11, 12).

The studies determining professionalism level and affecting factors in Ethiopia are limited to a few numbers with a clear recommendation for further investigation of factors influencing professionalism in nursing. Moreover to the best of our knowledge there is no study conducted in Hadiya zone regarding with professionalism in nursing & associated factors. Therefore this study was assessed professionalism and associated factors among nurses working in Hadiya Zone public hospitals. The outcome of this study helps Public hospitals and the FMOH in drafting policies and guiding principles of nursing professionalism in Ethiopia as well as for nursing professionals to confirm their status of professionalism.

## Method and materials

### Study design and period

Institution based concurrent mixed cross sectional study was conducted From May 09 to June 09, 2022, among nurses working in Hadiya zone public hospitals, south Ethiopia. For qualitative study Phenomenological study design was used.

### Study setting

Hadiya zone is one of the populated zones and located 230 kms away from Addis Ababa capital of Ethiopia and Hossana is its capital city. Administratively, the Hadiya Zone was organized by 4 administrative towns, 17 woreda, 305 rural Kebeles, and 30 urban Kebeles. Among five public hospitals namely, Wachamo Nigist Ellen Mohammed memorial comprehensive specialized teaching hospital, Shone, Bonosha, Gibe and Gimbicho primary hospitals; each of them has 316, 72, 58, 66 and 64 number of nurses respectively. Total numbers of nurses found in five hospitals were 576.

### Inclusion criteria and Exclusion criteria

For quantitative study all selected nurses on active duty during the study period were included in this study. For qualitative study nurses who have position in the hospital. Nurses who were in annual and maternal leave were excluded from this study.

### Sample size determination

The sample size was determined by using single population proportion formula using a basic assumption of 95% confidence level, 5% margin of error by considering the proportion of 33.4% for the level of professionalism in nursing with a study conducted in Jimma zone, Oromia region, Ethiopia (11).

### Sample size determination for qualitative part

For in-depth interview the numbers of participants were decided based on the level of data saturation. In the study saturation was obtained with six (6) participants.

### Sampling technique

Study participants were taken proportionately from each hospital. To select a study unit computer generated simple random sampling technique was used for quantitative study. For qualitative study purposive sampling technique was employed to select study participants (Key Informants) for in-depth interview. The Key informants for this study were the nurses who have a position in the hospital.

### Operational definitions

#### Professionalism

Adherence in all roles including practice standards, qualities, values and attitudes as stated in registered nurses association of Canada best practice guidelines (13).

#### High professionalism in nursing

participants who scored between 80 and 100% (136–170 points) of the total sum of the nursing professionalism scores (4).

#### Moderate professionalism in nursing

participants who scored 60–79% (102–135 points) of the total sum of perceived nursing professionalism scores

#### Low professionalism in nursing

participants who scored less than 60% (34-101 points) of the total sum of perceived nursing professionalism scores (4)

#### Poor organizational culture

Below half proportion of total score of organizational culture

#### Good organizational culture

Above half proportion of total score of organizational culture.

### Data collection instrument and procedure

The data was collected by self-administered questionnaire which has four parts. **Part I** socio demographic variables consisted of 9 items to collect information of sample characteristics.

**Part II:** The measurements of professionalism in Nursing, which was Self -administered professionalism assessment scale. The questionnaire was adapted from RNAO guideline which was validated and used in our country by Atsede F et al. consisted of 34 items related to self-appraisal scale to measure professionalism in nursing among nurses working in Hadiya zone Public hospitals whose Cronbach **α =** 0.95 (12).

The professionalism assessment self-appraisal scale (Likert) responses allow subjects to assess their level of professionalism by giving their agreement on a continuum (Strongly disagree=1, disagree=2, neutral=3, agree=4, and strongly agree=5).

**Part III:** Consisted of 8 items related to personal characteristics. From which 3 items were self-appraisal scale (Likert) responses to assess personal sense of accomplishment, the rests were dichotomous (yes or no) questions to assess personal related characteristics which are adapted from related literatures (11, 14).

**Part IV:** Measures organizational cultures which were adapted from Nursing Assessment Survey (NAS) used by Solomon et al. in Ethiopia and have 14 items in five subscales which contains five point Likert scale ranged from strongly disagree (1) to strongly agree (5) α=0.838 (11, 14).

Data collection was facilitated by five BSc nursing professionals with one supervisor. Principal investigator was made sudden observations during the data collection process. Training for data collection facilitators and supervisor was given one-day prior to data collection. Training was focused on purposes of study, the significance and appropriate meanings of each question.

For qualitative data in-depth interview with open ended questions was conducted. The data in in-depth interview (IDI) was collected by the principal investigator and each interview was held through recording audio after obtaining voluntary consents from interview participants. Data collection and probing of ideas was continued to the point of saturation.

### Data Quality Assurance

Pre-test was conducted on data collection tools on 5% (19) of sample size in Werabe comprehensive specialized hospital; in it clarity, logical sequence of questions and the total time it takes to finish the questionnaire were assessed before starting actual data collection. Each data collection facilitator was checked the questionnaires for completeness before winding up their visit to each study participant. Every night each questionnaire was reviewed by supervisor and the Principal investigator to check for completeness.

Identifying and describing the study participants based on their position was undertaken as a method of ensuring the data quality for qualitative study.

### Data Processing and Analysis

After data collection, each questionnaire was coded and entered into Epi data version 4.6 and exported to SPSS version 25 software package for the analysis of quantitative data. Descriptive statistics like frequencies, percentages, means and SDs were used to describe study subjects. The result was presented in the form of tables, figures and text using frequencies.

Ordinal logistic regression model was used to identify the effect of explanatory variables on outcome variable. Pearson and deviance goodness of fit test were conducted to check the model fitness. In which identified “***P”*** values were **0.550** and **0.997** respectively. Test of parallel lines was also checked for the fitness of assumptions; in the test ***P*** value of **0.571** was identified. Multicollinearity was checked by using VIF and tolerance test; their test values were VIF < 10 and T > 0.1. A bivariable and multivariable logistic regressions were used to identify the association of the independent variables with the dependent variable. Each variable which have p-value less than 0.25 at bivariable analysis was added in to the final model. Variables which have a p-value < 0.05 in the final model were declared statistically significant.

For qualitative data primarily, audio recorded data was heard repeatedly until the principal investigator became intimately familiar with the contents. The audio taped qualitative data was transcribed. After that it was translated in to English language. Then, codes or terms was identified and tallied through QDA minor lite software to come up with some categories, which later used to establish themes based on the objective of the study. Finally, thematic analysis was done and the finding was triangulated with the quantitative one.

### Ethical Consideration

Ethical approval for the study was obtained from the research ethics review committee of Bahir Dar University, College of Medicine and Health Sciences. Then, letters was written from Hadiya Zone Health bureau to the each public hospital in the zone. Participants were informed about the objectives of study, and written consent to participate them was requested. Participants were also informed that any information they provided was confidential and that the final report will not include personally identifiable information. Any participants who hadn’t willing to participate in the study were not forced to participate.

## Results

### Socio demographic Characteristics of study participants

From the total of 376 sample size 360 nurses were participated in this study with a response rate of 95.7%. The higher proportion of respondents were in the age range of 18-29 accounts 257 (71.4%). The higher proportion of the study participants, 201(55.8%) were from the Wachamo University Comprehensive specialized Hospital followed by Shone primary hospital 44 (12.2%), Gibe primary hospital 39 (10.8%) and Bonosha primary hospital 35 (9.7%). In the study higher proportion 190 (52.8%) were female participants and 197 (54.2%) were single. The majority of the respondents, 233 (64.7%), were degree holders and 333 (92.5%) were staff nurses in their working Unit. 6357.62 ± 940.473 was the mean monthly salary of the respondents. About 291 (80.8%) of the participants, had nursing experience of < 5 years. Among the total participants, 251 (69.7%) had no membership in the professional organizations, and 232 (64.4%) responded that they had not taken short term training and 278 (77.2%) had no experience of engagement in informal learning while only 82 (22.8%) had experience of E-learning. (Table 2)

**Table 1:** Socio-demographic Characteristics of Nurses Working in Hadiya Zone Public Hospitals, South Ethiopia 2022 (N=360)

| Characteristics | Categories | Frequency | Percent (%) |
| --- | --- | --- | --- |
| Age | 18-29 | 257 | 71.4 |
|  | 30-39 | 101 | 28 |
|  | ≥ 40 | 2 | 0.6 |
| Sex | Male | 170 | 47.2 |
|  | Female | 190 | 52.8 |
| Salary (mean± SD) |  | 6357.62 ± 940.473 |  |
| Marital status | Single | 197 | 54.7 |
|  | Married | 163 | 45.3 |
| Professional qualification | Diploma | 125 | 34.7 |
|  | BSc | 233 | 64.7 |
|  | MSc | 2 | 0.6 |
| College of completion | Private | 65 | 18.1 |
|  | Government | 295 | 81.9 |
| Participating in professional O. | Yes | 109 | 30.3 |
|  | No | 251 | 69.7 |
| Position at work | Head of department | 19 | 5.3 |
|  | Matron | 3 | 0.8 |
|  | Liaison officer | 5 | 1.4 |
|  | Staff nurse | 333 | 92.5 |
| Year of experience in nursing | <5 Years | 291 | 80.8 |
|  | 5 - 10 | 49 | 13.6 |
|  | >10 | 20 | 5.6 |
| Short term training | Yes | 128 | 35.6 |
|  | No | 232 | 64.4 |
| E- learning | Yes | 82 | 22.8 |
|  | No | 278 | 77.2 |

**Table 2:**
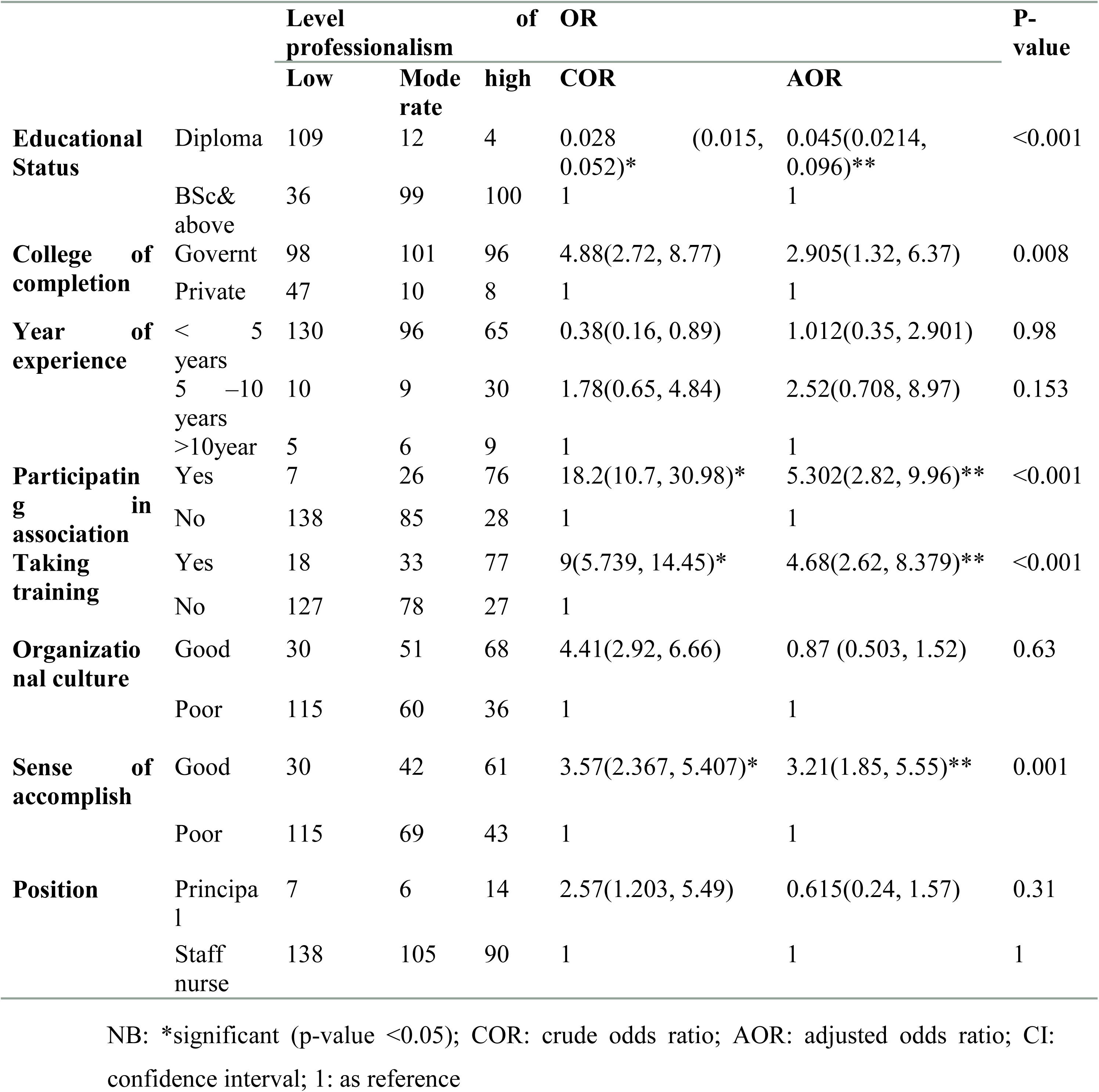
factors associated with professionalism in nursing in multivariable analysis among nurses working in Hadiya Zone Public Hospitals, South Ethiopia, 2022.

**Table 3:** Overview of categories (n=3) and subcategories (n=8)

| Major themes | Sub themes | Examples of subtheme |
| --- | --- | --- |
| **Governmental related factors** | Salary and incentives | **KI6** "... This is not in line with the Salary we receive". |
|  | Undermining nursing profession | **KI6** "... This resulted from undermined attitude of government and MOH for nursing profession" |
| **Individual related factors** | Attitude of nurses | **KI5** "... when nurses have a positive attitude toward their work" |
|  | Educational level | **KI2** "Your professionalism also rises as your educational level does" **KI4** "Education opportunity for nurse and a midwife is limited in this hospital" |
|  | Accepting profession | **KI6** "... You must first accept and love your vocation" |
|  | Participating in professional associations | **KI1** "... Participating in the associations has many advantages, for example it makes nurses to have a common goal" |
| **Institutional related factors** | Lack of adequate facility | **KI3** "... But in this hospital there is lack of facility to give appropriate service for patients". |
|  | Lack of training | **KI6** "... You must take part in many forms of training in order to build your professionalism" **KI4** "... but no training in this hospital" |

### Sense of Accomplishment

From the total respondents more than half of the participants 227 (63.1%) had a Poor personal sense of accomplishment. Among the study subjects about 95 (26.4%) and 105 (29.2%) participants were disagree with the idea of “I have opportunities to improve my skills and talents”; “I have opportunities to feel proud my work” respectively. (Figure 1)

**Figure 1:**
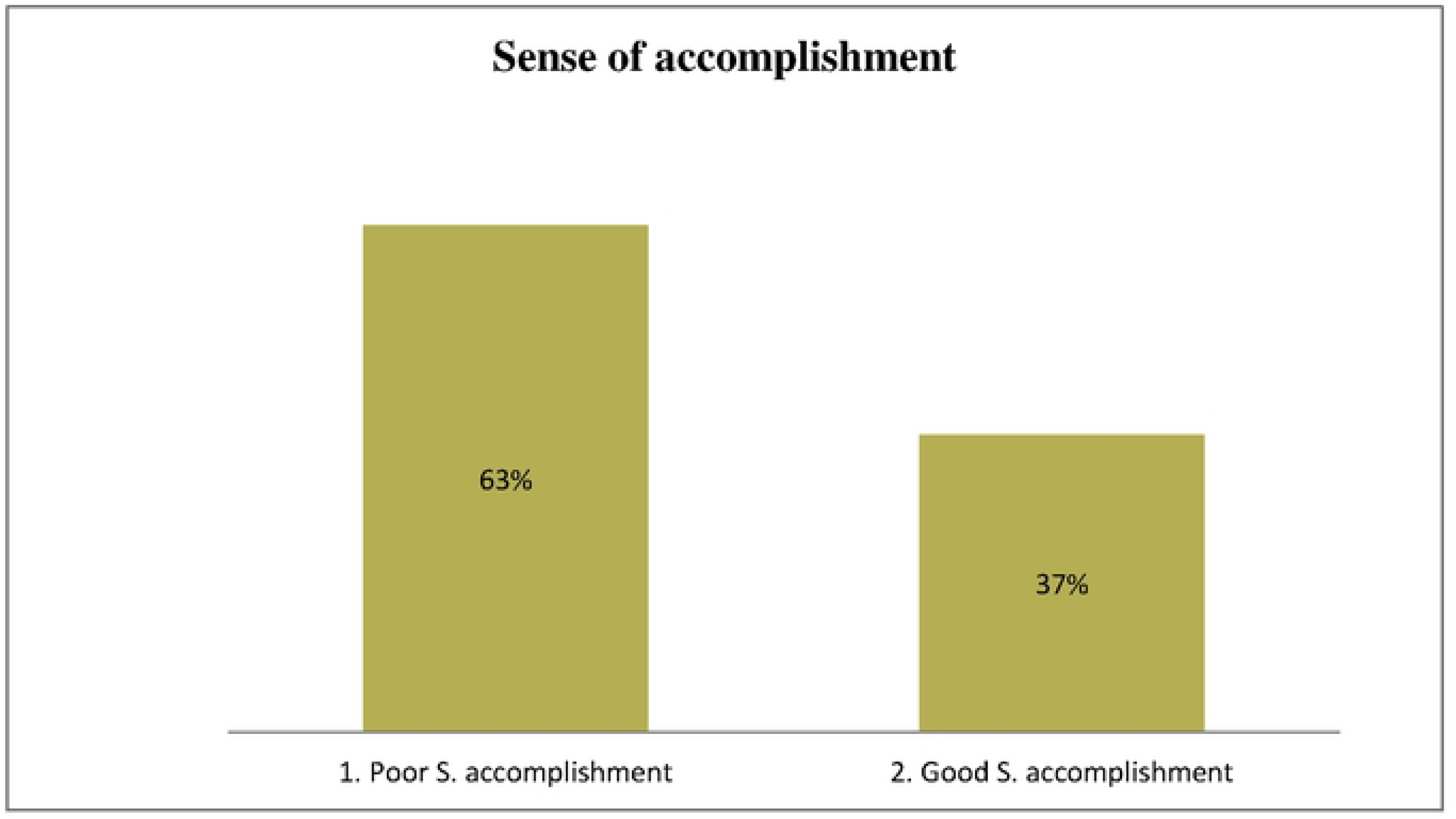
Sense of Accomplishment for nurses working in Hadiya zone Public Hospitals, South Ethiopia, 2022 (N= 360)

### Organizational culture

In the organizational culture assessment highest median score was in the area of strength of hospital cultures 8 ± 3 and followed by Recognition 7 ± 4, Affiliation 7 ± 4, Accomplishment 7 ± 4 and Power 5 ± 2. In the organizational culture assessment 211 (58.6%) of the respondent fail to share the organizations value. Figure 2

**Figure 2:**
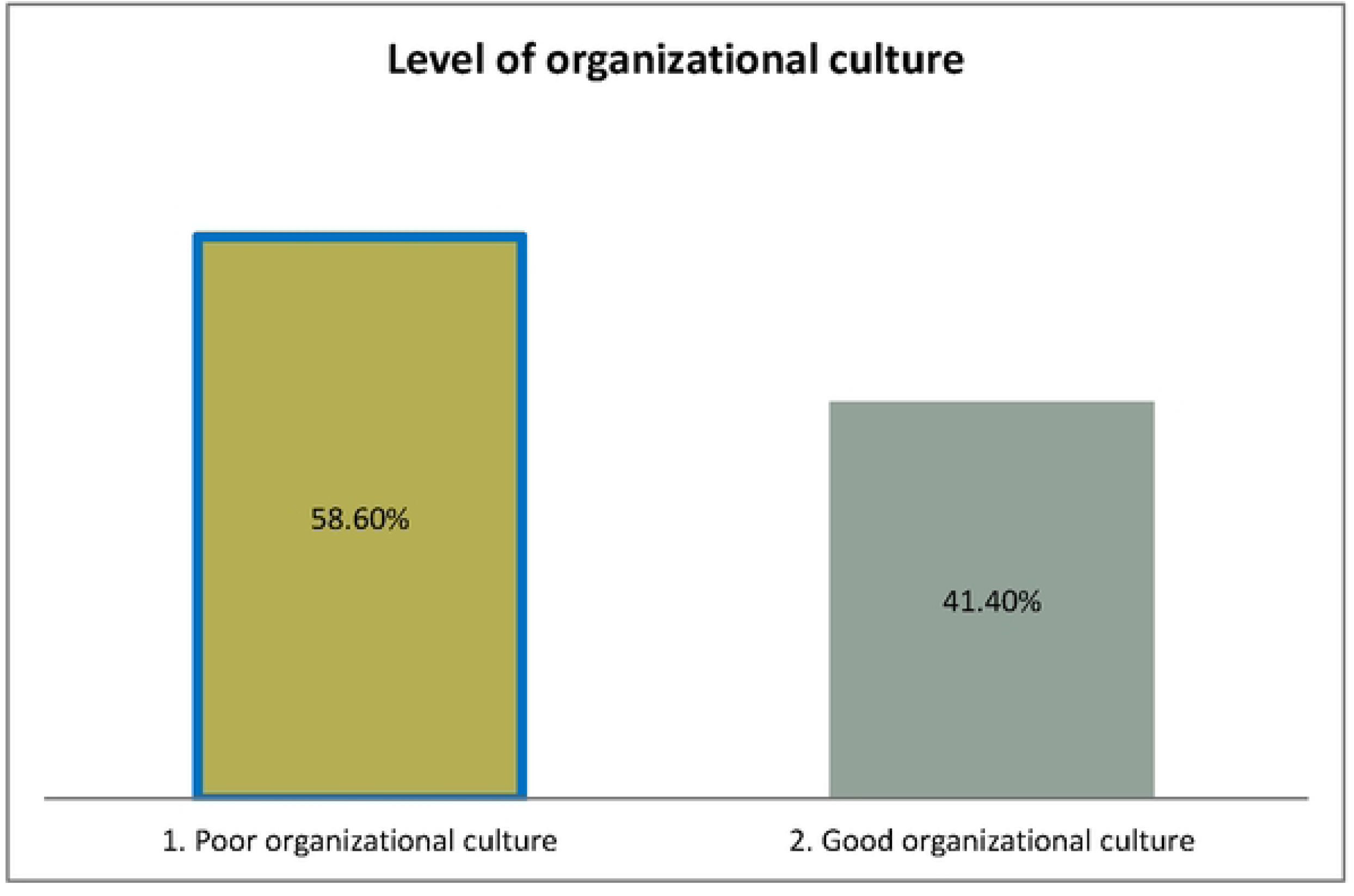
Level of Organizational culture in Hadiya zone Public Hospitals, South Ethiopia, 2022 (N= 360)

### Level of Professionalism

The median scores for the nurses in Hadiya zone public hospitals on the professionalism were 117.5 ± 60.75. The findings showed the respondents score highest on Ethics and value (22.00), followed by knowledge (21.0), Accountability (17.10), Spirit of inquiry (14.00), advocacy (12.00), collaboration and collegiality (11.00), Innovation and visionary (10.00) and Autonomy (8.00). From the professionalism score range; 145 (40.3%) of the respondents scored low professionalism; value ranging from 34-101 (Figure 3)

**Figure 3:**
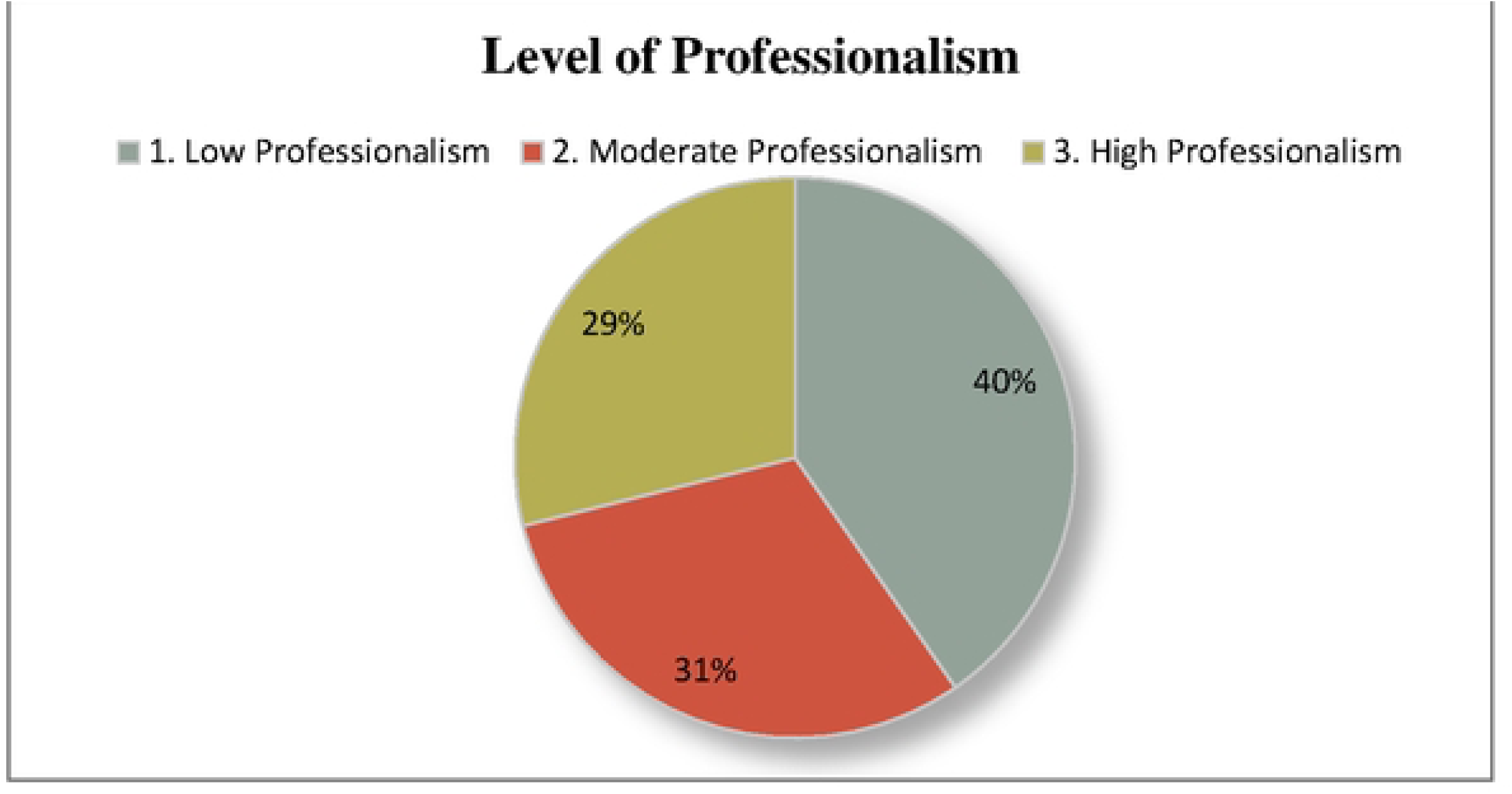
Level of professionalism among nurses working in Hadiya zone public hospitals South Ethiopia, 2022 (N= 360)

### Factors associated with Professionalism

Fourteen (14) independent variables such as, Age, Sex, Marital status, Current salary, Work setting, Year of experience in nursing profession; Educational level, Membership in professional organization, Organizational culture, College of completion, Working position in hospital, Taking training, E-learning experience and Personal sense of accomplishment were entered independently to see their independent effect on the level of professionalism among nurses working in Hadiya zone public hospitals.

Variables with p-valve ≤ 0.25 in Bivariable analysis were entered in the final model. In the model, Level of education, college of completion, personal sense of accomplishment, taking training, and participating in professional organizations were found to be significantly associated with the level of professionalism among nurses working in Hadiya Zone public hospitals.

In this study level of education was significantly associated with the professionalism in nursing. The odd of being in a higher category of professionalism was 0.045 times lower than for those who have professional qualification of diploma as compared to those who have BSc and above (AOR= 0.045, 95%CI; 0.021, 0.096). Nurses having BSc and above have 95% probability of falling on higher category of professionalism.

College of completion was the second factor that was significantly associated with professionalism in current study. The odd of being in higher category of professionalism was 2.9 times higher for the nurses who have completed their education in governmental college than Private. In this study college of completion can explains 18% variance of professionalism in nursing (AOR= 2.905, 95%CI; 1.32, 6.37).

Personal sense of accomplishment was also another significant explanatory variable for professionalism in nursing and it explains 11% of variance of professionalism in nursing in bivariate analysis. The odd of being in higher category of professionalism was 3.2 times higher for the nurses who have good sense of accomplishment than the nurses who had poor sense of accomplishment (AOR= 3.21, 95%CI; 1.85, 5.55)

Being a member of professional organization was found to have a positive association with professionalism among nurses working in Hadiya zone public hospitals and it explains 40% of the variance in nursing professionalism. the odds of being on higher professionalism category was 5.3 times higher for the nurses who are member for professional organization as compared to not being a member for professional organizations with 84% probability of falling on higher category of professionalism (AOR= 5.302 95%CI; 2.82, 9.96).

The last variable found to have a positive association was taking training. The odds of being on higher category of professionalism was nearly 4.7 times higher for the respondents who had ever take training than didn’t take training (AOR = 4.68, 95% CI, 2.62, 8.379). Those nurse who had ever take training have 82% probability of falling on higher category of professionalism.

### Result of Qualitative study

#### Socio demographic characteristics of key informants

In-depth Interviews were conducted from June 05-07/2022. Thematic saturation was obtained with six participants. Participants included WUCSH nurses’ leader or matron (n=1, 16.67%), WUCSH surgical ward head (n=1, 16.67%), Shone primary hospital matron (n=1, 16.67%), shone primary hospital medical ward head (n=1, 16.67%), Gimbicho primary hospital matron (n=1, 33.3%) and Gibe primary hospital emergency department head (n=1, 16.67%). A total of 5 males and 1 female were key informants in this study. In total, 3 major themes such as Governmental related factors; Individuals related factors and Institutional related factors were extracted by analyzing the data.

### Governmental related factors

Governmental related factors considered as external influencing factors of professionalism. That can influence externally to the individual’s behavior.

#### Salary and incentives

According to key informants, salary and incentive issues influenced professionalism. They claim that there is an imbalance between what they earn and what they sacrifice. As stated by KI6 *“…The nursing profession is all-encompassing; I am happy with it, but there are certain difficulties and Ethiopia is really boring. The amount of time you put in and the amount they pay for you are not proportional. Does the doctor spend time with his or her patients? Does specialist? Neither of them! Nurses, however, respect the patient’s time, act ethically, and provide the right treatment as needed. This is not in line with the Salary we receive”*.

#### Undermining nursing profession

As interviewers responded there is no focus, no proper recognition and acknowledgement from Ethiopian government for what nurses scarifying. As implied by key informants government and federal MOH undermine nursing profession. As stated by KI1….”*Nursing profession is undermined by MOH and governmental bodies. In the case of salary why those anesthetists and public health officers earns higher salary than nurses, what is the difference? The year of education is the same, is it for the job description? I don’t think so; it shows the undermining nursing profession”*.

Similarly as KI6 statement, “… *if mistake is made by nurse and physician, the only nurse is accountable for that mistake. This resulted from undermined attitude of government and MOH for nursing profession. Ethiopian government does not know about nursing profession properly. I think MOH also didn’t understand the problems that suffer nurses when compared to other professionals”*.

### Individuals related factors

Individual factors considered as inner processes which can be described as a set of values, attitudes, beliefs, and capabilities that are internal to the individual (15). In this study individual related factors were raised in the key informant interviews.

#### Attitude of nurses towards their profession

The attitude of nurses plays a significant impact in professionalism, according to key informants. In line with KI5’s argument professionalism will emerge when nurses have a positive attitude toward their work and exhibit good patient care. *“… In my opinion, professionalism will emerge when nurses have a positive attitude toward their work,” the author says”*.

Another key informant (KI1) makes the suggestion that nurses who have a bad attitude toward their profession will stop practicing. *“If a person’s attitude about a certain profession changes, they may not stay in that profession and won’t be able to advance their professional skills. Therefore, it should be necessary to focus on changing the nurses, governments and nurses’ managers’ attitudes”*.

#### Accepting profession

The characteristics that can impact nursing professionalism were acceptance and love of one’s work. Failure to love and embrace one’s own profession is a major cause of resignation or turnover. According to KI6 it’s crucial to accept and love one’s profession if you want greater outcome *“… You must first accept and love your vocation; next, you must understand its importance and work diligently in accordance with it; and finally, you must be knowledgeable and skilled in your field. The other is looking for and coming up with ideas to advance your career”*.

#### Educational status

Upgrading educational level is regarded as one of the fundamental requirements for nursing professionalism, the interviewer explained. According to respondents’ explanations, a higher educational degree has a positive impact on professionalism. As KI2’s stated, *“Upgrading educational status can enhance a nurse’s professionalism. Your professionalism also rises as your educational level does—from a diploma to a degree, a master’s to a PhD, and so forth”*.

As indicated by key informants KI1 and KI4 despite upgrading educational level has direct positive impact on nursing professionalism, there is a limited opportunities to upgrade educational level in nursing *“… When you become more advanced, you will carry out research and learn new things, which will subsequently be applied to improving nursing curriculum and education. The nursing curriculum, for instance, are based on early researches that were undertaken, but few nursing practitioners actually accomplish this.* **As explained by KI4** “… *In this hospital the chance to get education opportunity for nurse and a midwife is limited in to a few numbers”*.

#### Participating in professional organizations

participating in professional association gives a sense of belongingness for them. It promotes collegiality, which can encourage mutual respect and trust. As stated by KI1: *“Participating in the associations has many advantages for example it create common sense among nurses to participate in research projects, in curriculum development and to challenge government to remedy some gaps dealing with compensation and incentives. Every nursing-related topic in Ethiopia, such as queries about government, meetings, and research-related matters, is better handled through the Ethiopian Nursing Association”*.

### Institutional related factors

This theme recognizes the broader workplace conditions in which the phenomenon of professionalism exists and how professionalism is both influenced and constrained by these institutional contexts. In this study finding in adequate facility and training were raised as the institutional factors.

#### Inadequate facility

Lack of proper facilities in hospital settings, as described by key informants, will affect professionals’ skill and render nurses incompetent in practical skills and evidence-based practice. It will be challenging to provide patients with the necessary care when there are no facilities. Your professionalism can suffer if there aren’t adequate facilities to hold caring services, according to KI4. *“…In the absence of adequate facility, you can’t provide the standard of service that is expected from you. Your professionalism could be hampered by this situation. In the presence of a complete facility, you may provide your patients with suitable and amusing services, which will improve your skills but in this hospital there is lack of facility to give appropriate service for patients”*.

#### Training

According to key informants, training is crucial for nurses to grow professionally. Getting different types of training is an important to doing advanced and updated tasks, and it is crucial to expand one’s knowledge and skill set. As KI6 explains, *“… You must take part in many forms of training in order to build your professionalism. For instance, if you take part in emergency care training, you will be familiar with emergency care in depth.* As stated by KI4*: “Creating a proper environment and facilitating the requirements, allows for professional development, however there is no training for nurses and midwives in this hospital.”*

## Discussion

The purpose of this study was to assess the professionalism of nurses working in Hadiya zone public hospitals, as well as the associated factors. The study’s findings showed that 145 nurses (40.3%) had low levels of professionalism, with a 95% confidence interval (35.2% -45.5%). The level of nursing professionalism was found to be significantly influenced by a variety of factors. These included a Sense of accomplishment, participating in professional organization, taking up-to-date Training, Education levels, and College of completion.

The results of the current study showed that the professionalism of nurses working in Hadiya zone public hospitals was low. This was consistent with the study done in Jimma public hospitals. Other studies also are in line with current study finding. The report of the studies which conducted among Turkish and Japanese nurses also revealed that low levels of professionalism (10, 11, 16). On the other hand, the finding of this study was low when compared to a study conducted in Iran and USA reported that a moderate and high levels of professionalism among nurses respectively (17, 18).

The possible reason for the discrepancy could be low professional development, excessive workload, long working hours, and insufficient services supplied in the nursing profession in developing nations like Ethiopia (4). For example in the case of workload; nurse to patient ratio in Ethiopia ranges from **1:6 to 1:12** based on the individual institution patient load and nurse availability (19). But in USA the nurse to patient ratio is ranges from 1:1 to 1:6 (20) this could makes difference and have effect on the nurses professionalism level (4).

The other possible reasons could be the negative factors that can hinder professionalism in nursing such as, perception in the community about nursing as a profession, the hierarchic structures of hospitals, the focus of nursing on tasks, a lack of personnel and equipment, low salaries, and weakness in organized labor (5). The health care delivery system may also be another implication for the discrepancy of study’s findings which can be heavily influenced by Social, technological, and economic development of the countries this can have an impact on nurse’s professionalism; especially quick changes in the health sector have a pronounced impact on the demand for professional nurses (3).

Educational level was found to be a strongly associated with nursing professionalism in this study. When comparing nurses with diploma holders to those with BSc and higher degrees of education, the odds of being in a higher category of professionalism was 0.045 times lower. This finding was congruent with qualitative study finding. Upgrading educational level is regarded as one of the fundamental requirements for nursing professionalism, the interviewer explained. According to respondents’ explanations, a higher educational degree has a positive impact on professionalism. As KI2’s stated, *“Upgrading educational status can enhance a nurse’s professionalism. Your professionalism also rises as your educational level does—from a diploma to a degree, a master’s to a PhD, and so forth”*.

This may be explained by the fact that study participants with a BSc or higher in nursing are more aware of the updated information, and difficulties in nursing-related issues that support professional development. To be considered a professional in their area today, several disciplines, including nursing, require graduates to hold a degree at the BSc level. The studies done in Turkey and Jimma supported this finding (5, 11).

The second factor in the current study that predicts professionalism was participating in a professional organization. In the study, nurses who were members of professional organizations had, on average, 5.3 times higher odds of falling into a higher professionalism category than those who were not. This finding was supported by the qualitative study finding. According to key informant response, Participating in professional associations gives nurses a sense of belonging and encourages collegiality. As stated by KI1: *“… Participating in the associations has many advantages, for example it makes nurses to have a common goal through it to participate in research projects, in curriculum development and to challenge government to remedy some gaps”*.

Possible reason might be joining professional groups can help with career development, keeping up with the trends issues, increasing skill development, and providing possibilities for professional networking. Professional associations offer a crucial way to deal with complex challenges that individual members cannot solve. Nursing association also play a key role in assisting nurses in developing their professionalism (21). The finding was supported by the study conducted in USA (18).

On the other hand, Personal sense of accomplishment was one of the study’s predictive variables. For nurses with a good sense of accomplishment compared to nurses with a low sense of accomplishment, the odds of being in a higher category of professionalism were on average 3.2 times greater. This suggests that drive and success are indicators of one’s sense of accomplishment. Better results for nurses, hospitals, and the patients they serve will increase personal satisfaction (22).

Another factor that was significantly associated with nursing professionalism was having updated training. For nurses who had taken updated training compared to nurses who hadn’t, the odd of being in a higher category of professionalism was, nearly 4.6 times greater. This finding was congruent with the qualitative study finding. This could be attributed that taking updated training and courses refresh and develop nurse’s practical skills and knowledge levels. Providing qualified health care for patients requires up-to-dated knowledge and expertise, professional performance, essential management skills, and the ability to provide safe and appropriate legal and ethical services. In this context, taking updated training is essential. The research conducted in Mekelle supported this result (12).

College of completion was another positive indicator of nursing professionalism in current study. Finding of the study imply that the odds of being in higher category of professionalism was 2.9 times higher for those the nurses who were completed their education in Governmental College than nurses who were in private. This might be explained due to rapid and unplanned expansions of private college for the sake of opening access, without adequate infrastructure and resources are potentially dangerous for quality of education. Private colleges are on the challenge to balance poor funding, scarcity of qualified instructors, poor infrastructure, and poorly qualified students (23). This could have negative effect on the knowledge and skill level of nurses. To achieve professional status; to adhere with nursing care standard and to provide quality service, having strong powerful basis in theory, practice, and professional education in nursing discipline is needed because they are a basis to establish professionalism among nurses (3)

## Limitation of the study

Limitation for this study was asking the respondents to determine their perceived level of professionalism by self –report, this is not best method to identify level of professionalism. So, it should be considered for future study to measure professionalism. Second limitation, Professionalism was assessed by using only attitudinal components. Better to assess attitudinal components alongside with behavioral components of professionalism in nursing.

## Conclusion

The results of this study showed that nurses’ levels of professionalism were low, and the significant factors that contributed to this were; level of education, taking training, participating in professional organizations, sense of accomplishment and college of completion. Salary and incentives, lack of training, Accepting one’s own profession, upgrading level of education, Undermining nursing profession from government and MOH, lack of facility, attitude of nurses toward nursing profession and participating in professional associations were the influencing factors of professionalism in nursing as described by key informants.

## Recommendations

1. Nurses working in this area better to actively participate in Ethiopian nursing association and in other health related professionals association.
2. FMOH should give the equal value for nursing as the others profession.
3. Pure qualitative studies should be considered to explore further barriers of professionalism in nursing.

## Ethical Approval

The study obtained ethical clearance from the Bahirdar University Institutional Review Board (Ref. No-412/2022). Written permission for data collection was provided to the Hadiya zone Administrative Office, WUCSH, SPH, HPH, BPH, and GPH. The purpose and significance of the study were explained to the participants, and they were informed of their right to withdraw from the study at any time. To maintain the privacy and confidentiality of the participants, no personal identification such as names was collected. Verbal informed consent was obtained from each study participant before data collection.

## Authors’ Contributions

The study was conceptualized and the research tool was developed by Tadele Mengesha, Beriso Furo and Ephrem Mamuye. They also conducted the research, carried out the statistical analysis, and wrote the manuscript. Tadele Mengesha and Mitiku Abera participated in the study design and tool development, performed statistical analysis, and contributed to drafting the manuscript. All authors read and approved the final manuscript.

## Data Availability

The corresponding author will provide data upon request Address of corresponding author: Phone number: +251 916227210

## Acknowledgments

The authors express their gratitude to the Bahirdar University Institute of Health and Department of Nursing, for their guidance and support in the preparation of this research thesis. They also extend their thanks to WUCSH, SPH, HPH, BPH, and GPH for their cooperation. The authors would also like to acknowledge the data collectors, study participants, and supervisors for their valuable contributions to the study.

## Availability of data

The corresponding author will provide data upon request

Address of corresponding author

Phone number: +251 916227210

## LIST OF ABBREVIATIONS AND ACRONOMYS

ANA: American Nurses Association
AOR: Adjusted Odd Ratio
BPH: Bonosha Primary Hospital
BSc: Bachelor of Sciences
CI: Confidence Interval
COR: Crude Odd Ratio
ENA: Ethiopian Nurses Association
ERC: Ethical Review Committee
FMOH: Federal Ministry of Health
GPH: Gimbicho Primary Hospital
HPH: Homacho Primary Hospital
IDI: In Depth Interview
QDA: Qualitative Data Analysis
RNAO: Registered Nurses Association of Ontario
RNs: Registered Nurses
SPH: Shone Primary Hospital
SPSS: Statistical Packages for Social Sciences
SRS: Simple Random Sampling
WHO: World Health Organization
WUCSH: Wachamo university comprehensive specialized hospital

